# The BioNTech / Pfizer vaccine BNT162b2 induces class-switched SARS-CoV-2-specific plasma cells and potential memory B cells as well as IgG and IgA serum and IgG saliva antibodies upon the first immunization

**DOI:** 10.1101/2021.03.10.21252001

**Authors:** Anne S. Lixenfeld, Inga Künsting, Emily L. Martin, Vera von Kopylow, Selina Lehrian, Hanna B. Lunding, Jana S. Buhre, Janna Quack, Moritz Steinhaus, Tobias Graf, Marc Ehlers, Johann Rahmöller

## Abstract

To treat the SARS-CoV-2 virus, that enters the body through the respiratory tract, different vaccines in particular against the SARS-CoV-2 spike (S)-protein have been developed or are in the development process. For the BioNTech / Pfizer mRNA vaccine BNT162b2, which is injected twice, protection against COVID-19 has been described for the first weeks after the second vaccination. The underlying mechanisms of defense and the long-term effectiveness of this vaccine against COVID-19 are currently under investigation.

In addition to the induction of systemic antibodies (Abs), Ab responses in the respiratory tract would help to form a first line of defense against SARS-CoV-2. Furthermore, protection depends on Fab-part-dependent neutralizing capacities, however, Fc-part-mediated effector mechanisms might also be important. Long-term defense would be based on the induction of long-lived antibody-producing plasma cells (PCs) and memory B cells.

Here, we established different assays to analyze anti-SARS-CoV-2-S IgG and IgA Abs in blood serum and saliva as well as SARS-CoV-2-S1-reactive IgG and IgA PCs and potential memory B cells in the blood of individuals upon their first immunization with BNT162b2.

We show that the vaccine induces in particular anti-SARS-CoV-2-S IgG1 and IgG3 as well as IgA1 and in some individuals also IgG2 and IgA2 serum Abs. In the saliva, we found no anti-SARS-CoV-2-S IgA, but instead IgG Abs. Furthermore, we found SARS-CoV-2-S reactive IgG+ blood PCs and potential memory B cells as well as SARS-CoV-2-S reactive IgA+ PCs and/or potential memory B cells in some individuals.

Our data suggest that the vaccine induces a promising CD4+ T cell-dependent systemic IgG1 and IgG3 Ab response with IgG+ PCs and potential memory B cells. In addition to the systemic IgG response, the systemic IgA and saliva IgG response might help to improve a first line of defense in the respiratory tract against SARS-CoV-2 and its mutants.

## Introduction

SARS-CoV-2 (Severe acute respiratory syndrome coronavirus type 2), origin of Coronavirus Disease 2019 (COVID-19), has led to a global health threat. The virus infects cells in the respiratory tract through its transmembrane spike (S)-protein. The S protein contains a receptor binding domain (RBD) that interacts with the host’s membrane bound angiotensin-converting enzyme 2 (ACE2). A range of different vaccines against SARS-CoV-2 has been developed within just a few months, led by novel mRNA-based compositions such as the BioNTech / Pfizer vaccine BNT162b2 (Gaebler and Nussenzweig, 2020; Krammer, 2020). Here, modified mRNA coding for the viral spike-protein is embedded in lipid nanoparticles and injected intramuscular twice.

BNT162b2 appears to be safe and effective in producing SARS-CoV-2-specific neutralizing IgG antibodies (Abs) that inhibit the binding of the RBD to ACE2 (Wang et al., 2021) as well as drastically reduces the number of infected individuals in the weeks following the second immunization (Polack et al., 2020; Sahin et al., 2020). Furthermore, the vaccine can induce potential memory B cells (Wang et al., 2021).

However, it is not clear yet whether long-lived Ab secreting plasma cells (PCs), that survive longer than a few months, are induced by the vaccine and what the protective potential of the induced memory T and B cell and Ab responses is. In addition, not only the Fab-part-mediated neutralizing capabilities of Abs, but also their ability to crosstalk to other components of the immune system, determined by the Fc-part-dependent isotype and subclass might play an important role (Lilienthal et al., 2018). Different adjuvants induce qualitatively distinct immune responses (Bartsch et al., 2020) and it is open to question what kind of Ab isotypes and subclasses this mRNA lipid nanoparticle vaccine induces.

Furthermore, it is unclear whether the vaccine foremost generates a systemic serological immune response or a mucosal one in the respiratory tract as well. There is accumulating evidence for the critical importance of mucosal Abs in the defense against mucosal pathogens (Horton and Vidarsson, 2013; Krammer, 2020; Ruprecht et al., 2019; Staples et al., 2016). The respiratory system is the primary site of entry, multiplication and shedding of SARS-CoV-2 (Isho et al., 2020; Munster et al., 2020; Rockx et al., 2020), so mucosal Ab secretion after vaccination could play a beneficial role in preventing infection or at least reduce viral shedding. In this context, protecting mechanisms in the upper respiratory tract by especially secretory IgA1 and of IgG in the lower part of the respiratory tract have been described (Krammer, 2020).

Here, we analyzed anti-S1 (the extracellular part of S containing the RBD) and anti-S2 (the transmembrane and intracellular part of S) IgG and IgA Abs in the serum and saliva as well as S1-antigen-specific blood B cell responses of 19 individuals after the first immunization of the BNT162b2 vaccine to further characterize the B cell and Ab response.

## Material and Methods

### Vaccination study cohort and control individuals

Subjects were recruited at the University Hospital Center Schleswig-Holstein, Lübeck, Germany from December 2020 to February 2021: (i) 19 individuals injected with one dose (30 µg) of the BioNTech/Pfizer vaccine BNT162b2 and without known SARS-CoV-2infection history, (ii) one non-vaccinated individual without SARS-CoV-2 infection history as negative control and (iii) 9 non-vaccinated individuals with past (4) or current (5; of the intensive care unit (ICU) of the hospital) SARS-CoV-2infection as positive controls for antigen-specific cell staining and Ab detection (**Fig. 1 and Supplementary Table 1**). Additionally, negative saliva controls were collected from 18 individuals without known COVID-19 history (**Fig**.**1 and Supplementary Table 1**). Blood samples and saliva were collected after obtaining written informed consent under the local ethics board–approved protocols 19-019(A) and 20-123 (Ethics Committee of the University of Lübeck, Germany).

**Fig. 1.**
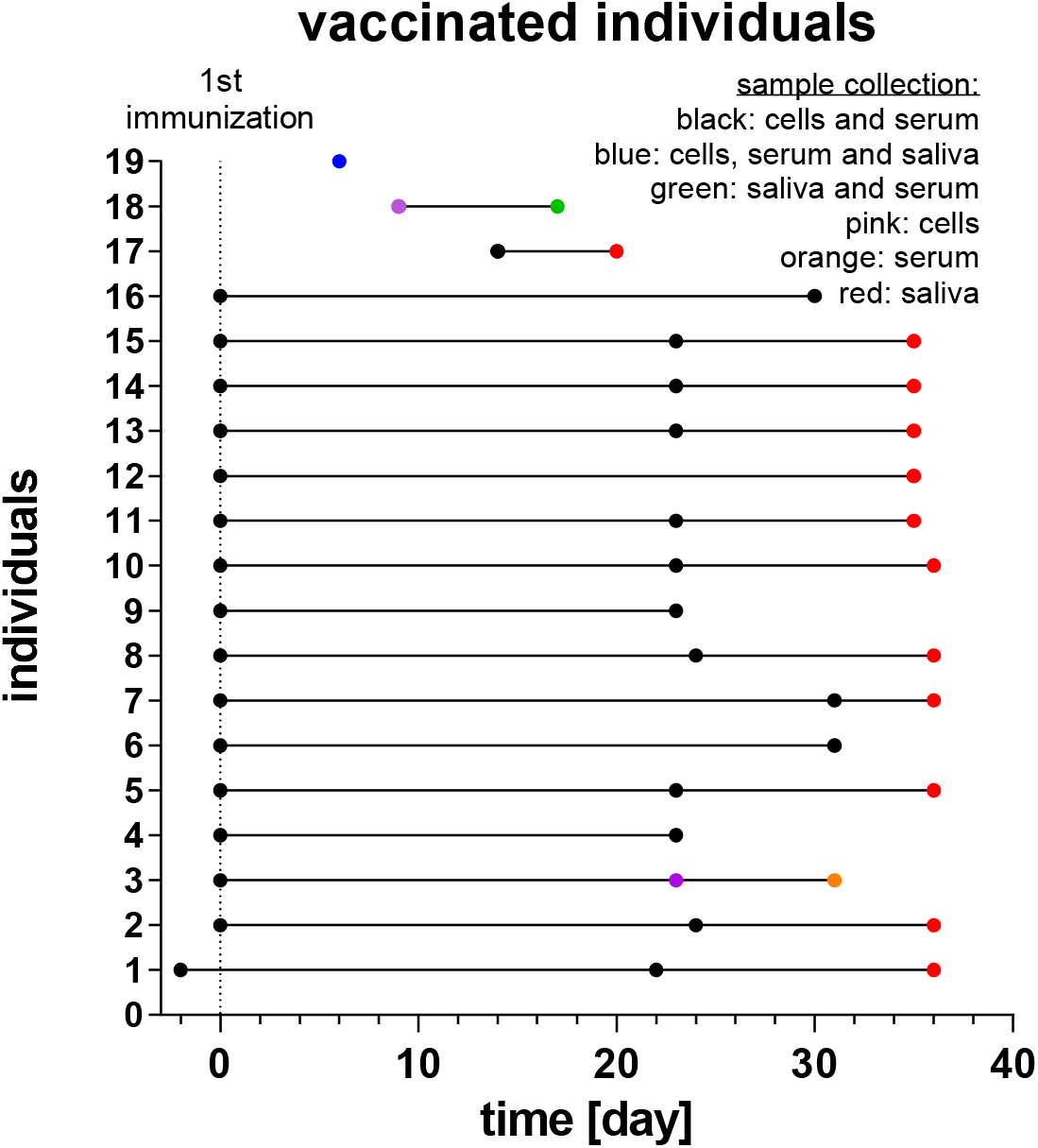
Subjects of the vaccination study cohort and time points of sampling. Individuals of the vaccination study cohort and the time points of blood cell, serum and saliva sampling.

### Blood serum and saliva antibody detection

Blood serum and saliva samples were collected for ELISA on the indicated days. Saliva was collected with the Saliva Collection system-ORACOL Plus S14 (Malvern Medical Developments, United Kingdom) and frozen before use. The EUROIMMUN SARS-CoV-2 S1 IgG (EUROIMMUN, Luebeck, Germany; #EI 2606-9601-2 G), the EUROIMMUN SARS-CoV-2 S1 IgA (#EI 2606-9601-2 A, containing their renewed buffer system), and the EUROIMMUN SARS-CoV-2-NCP IgG (#EI 2606-9601-2 G) ELISA were performed according to the manufacturer’s instructions.

Alternatively, 96-well ELISA plates were coated with 4 µg/ml of SARS-CoV-2-S1 (ACROBiosystems, Newark, DE 19711, USA; #S1N-C52H3) or -S2 (ACROBiosystems; #S2N-C52H5) antigen to identify anti-S1- and S2 Ab isotypes and subclasses (IgG, IgG1-4, IgA, IgA1, IgA2). The plates were washed with 0.05% Tween 20 in PBS to remove unbound antigen. In case of IgA, IgA1 and IgA2 serum as well as IgG and IgA saliva detection additional blocking was performed with 0.05% Tween 20, 3% BSA in PBS. Subsequently, serum (diluted 1/100 for IgG and IgG subclasses and 1/50 for IgA and IgA subclasses in 0.05% Tween 20, 3% BSA in PBS) or saliva (diluted 1/10) were added. Bound Abs were detected with horseradish peroxidase (HRP)-coupled polyclonal goat anti-human IgG Fc (#A80-104P) or IgA (#A80-102P)-specific Abs purchased from Bethyl Laboratories (Montgomery, TX, USA), or monoclonal anti-human IgG1 (clone HP-6001), IgG2 (clone HP-6014), IgG3 (clone HP-6050), IgG4 (clone HP-6025), IgA1 (clone B3506B4), IgA2 (clone A9604D2)-specific Abs purchased from Southern Biotech (Birmingham, AL, USA) in 0.05% Tween 20, 3% BSA in PBS. After incubation with the 3,3′,5,5′-tetramethylbenzidine (TMB) substrate (BD Biosciences, San Diego, CA, USA), the optical density (OD) was measured at 450 nm. To control secondary Ab specificity, ELISA plates were coated with 4 µg/ml of human monoclonal anti-collagen type VII IgG1-4, IgA1 or IgA2 Abs with identical V(D)J sequences (Recke et al., 2010 and 2014).

### Flow cytometric analysis

Blood samples were collected in EDTA-tubes and processed within the next 3 hours for flow cytometric analysis (Attune Nxt; Thermo Fisher Scientific) of different B cell populations. Peripheral blood mononuclear cells (PBMCs) were obtained by gradient centrifugation in biocoll. 20 million PBMCs were used for one staining. The following fluorochrome-coupled Abs were used for surface staining: anti-CD19 (Biolegend; clone HIB19), anti-CD38 (Biolegend: HIT2), anti-IgG-Fc (Biolegend; M1310G05) and anti-IgA (Miltenyi; IS11-8E10) as well as LIVE/DEAD Fixable Near-IR stain (Thermofisher; L34976). For additional intracellular staining, samples were fixed with Cytofix/Cytoperm according to the manufacturer’s instructions (BD Biosciences) followed by permeabilization (0.05% saponin, 0.1% BSA in 0.05 x PBS) and additional staining with anti-IgG and anti-IgA as well as SARS-CoV-2-S1 (biotin-coupled; Acro; #S1N-C82E8) and fluorochrome-coupled streptavidin (Biolegend). Flow Cytometry Standard (FCS) 3.0 files were analyzed with FlowJo software version X 0.7 (BD Biosciences).

### Statistical analysis

ELISA experiments were done twice. Statistical analyses were performed using GraphPad Prism v6.0 (GraphPad, La Jolla, CA). Data were presented as mean values ± SD. Differences between two groups were estimated with unpaired two-tailed Student’s test. P < 0.05 was considered to indicate a significant difference (*P < 0.05, **P < 0.01 and ***P < 0.001).

## Results

### Study group and controls

To investigate SARS-CoV-2-S1 and -S2-reactive blood B cells as well as IgG and IgA Abs in the serum and saliva after one time immunization with the BioNTech / Pfizer vaccine BNT162b2, 19 vaccinated subjects without COVID-19 history were investigated before and after the first (before the second) immunization as indicated (**Supplementary Table 1 and Fig. 1**). Additionally, samples of one non-vaccinated individual without known COVID-19 history and 9 non-vaccinated individuals with past (4) or current (5; of the intensive care unit (ICU) from the hospital) SARS-CoV-2 infection were used as negative or positive controls, respectively (**Table 1**). Further, saliva of 18 individuals without known COVID-19 history were collected and used as negative controls (**Table 1**).

### The vaccine induces SARS-CoV-2-S1 and -S2 reactive IgG1 and IgG3 as well as IgA1 serum Abs

Blood sera were collected up to two days before and at the indicated time point after the first immunization of the vaccination study cohort (**Supplementary Table 1 and Fig. 1**), as well as from individuals with past or current SARS-CoV-2 infection and analyzed with different commercially available anti-SARS-CoV-2 IgG ELISA from EUROIMMUN (anti-SARS-CoV-2-S1 IgG and anti-SARS-CoV-2-NCP (nucleocapsid protein) IgG) and anti-SARS-CoV-2-S1 and -S2 IgG ELISA established in our laboratory (HL-1 anti-SARS-CoV-2-S1 and –S2 IgG) (**Fig. 2A-C**). The results of the HL-1 anti-SARS-CoV-2-S1 IgG ELISA correlated well with the results of the EUROIMMUN anti-SARS-CoV-2-S1 IgG ELISA. (**Fig. 2D**). All except two vaccinated individuals showed detectable serum IgG Abs against SARS-CoV-2-S1 and –S2 (**Fig. 2A and C**). One of these two individuals showed no detectable specific IgG Abs even three weeks after vaccination (proband nr. 11). The first immunization of the other individual took place just six days before serum collection (proband nr. 19). All vaccinated individuals showed no increase of anti-NCP IgG Abs after vaccination (**Fig. 2B**).

**Fig. 2.**
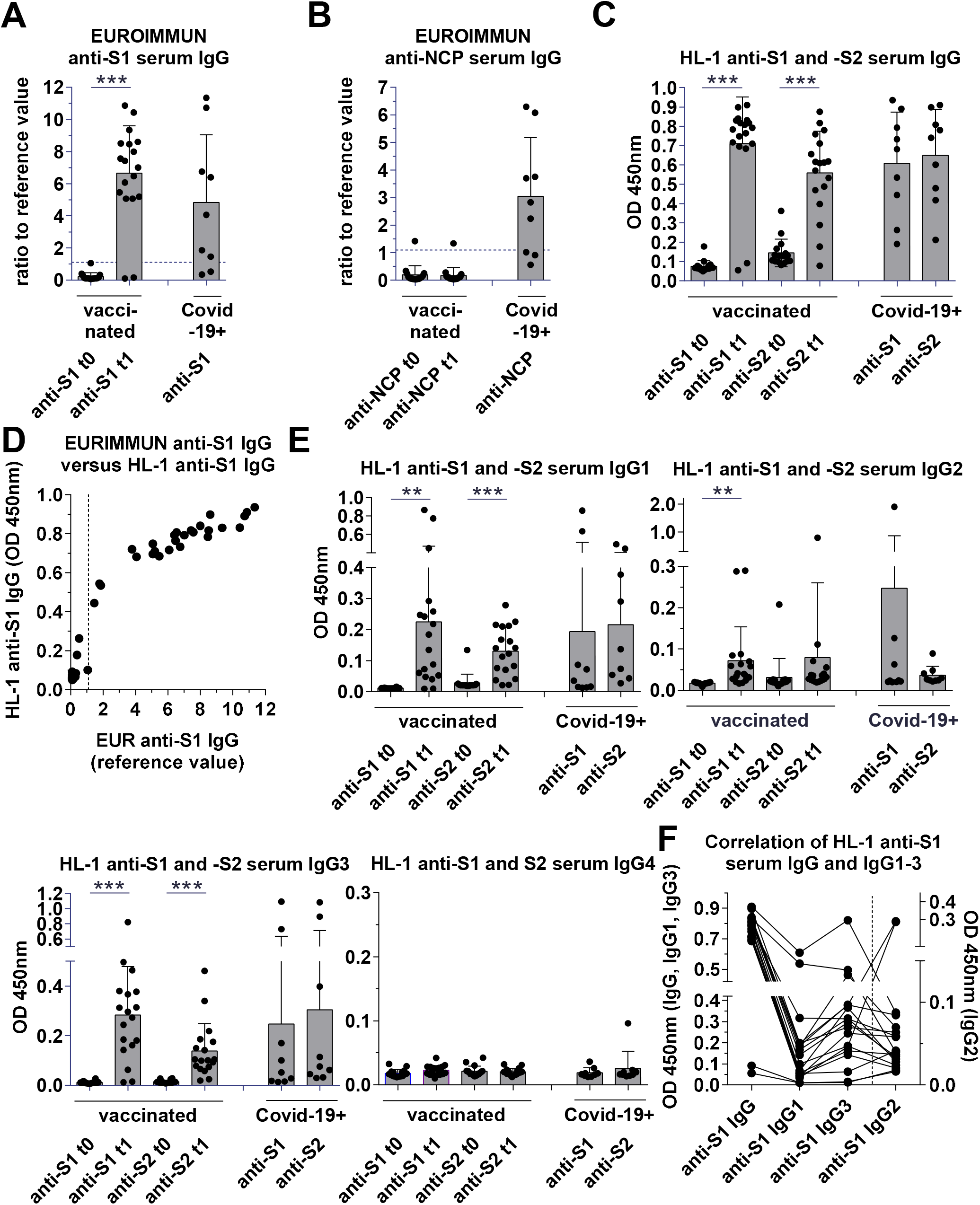
SARS-CoV-2-S1 and -S2 reactive serum IgG and IgG1-4 Ab ELISA. Sera were collected from vaccinated individuals before and after the first vaccination as well as non-vaccinated individuals with past or current COVID-19 infection and analyzed by anti-SARS-CoV-2-S1 and -S2 IgG and IgG1-4 ELISA. (**A**) EUROIMMUN anti-SARS-CoV-2-S1 IgG ELISA. (**B**) EUROIMMUN anti-SARS-CoV-2-NCP IgG ELISA. (**C**) HL-1 anti-SARS-CoV-2-S1 and -S2 IgG ELISA. (**D**) Correlation between the EUROIMMUN and the HL-1 anti-SARS-CoV-2-S1 IgG ELISA. (**E**) HL-1 anti-SARS-CoV-2-S1 IgG1-4 ELISA. (**F**) Correlation between HL-1 anti-SARS-CoV-2-S1 IgG and IgG1-3 ELISA results.

To analyze SARS-CoV-2-S1 and –S2 specific IgG1-4 subclass Abs, we used well established monoclonal anti-human IgG1-4 secondary Abs in our HL-1 ELISA system (**Supplementary Fig. 1**). The vaccinated IgG+ individuals developed anti-SARS-CoV-2-S1 and -S2 IgG1 as well as IgG3, but no IgG4 Abs (**Fig. 2E and F**). Some vaccinated individuals also developed anti-S1 and -S2 IgG2 Abs, in particular proband nr. 3 and 6 (**Fig. 2E and F**).

To analyze anti-SARS-CoV-2 serum IgA Abs we used the EUROIMMUN anti-SARS-CoV-2-S1 IgA ELISA kit and established an anti-SARS-CoV-2-S1 and -S2 IgA ELISA in our laboratory (named HL-2 anti-SARS-CoV-2-S1 and -S2 IgA) (**Fig. 3A and B**). The results of the HL-2 SARS-CoV-2-S1 IgA ELISA correlated well with the EUROIMMUN SARS-CoV-2-S1 IgA ELISA (**Fig. 3C**). All vaccinated individuals except of the two mentioned individuals (no. 11 and no. 19) showed detectable serum IgA Abs against SARS-CoV-2-S1, however, with highly different levels (**Fig. 3A and B**),

**Fig. 3.**
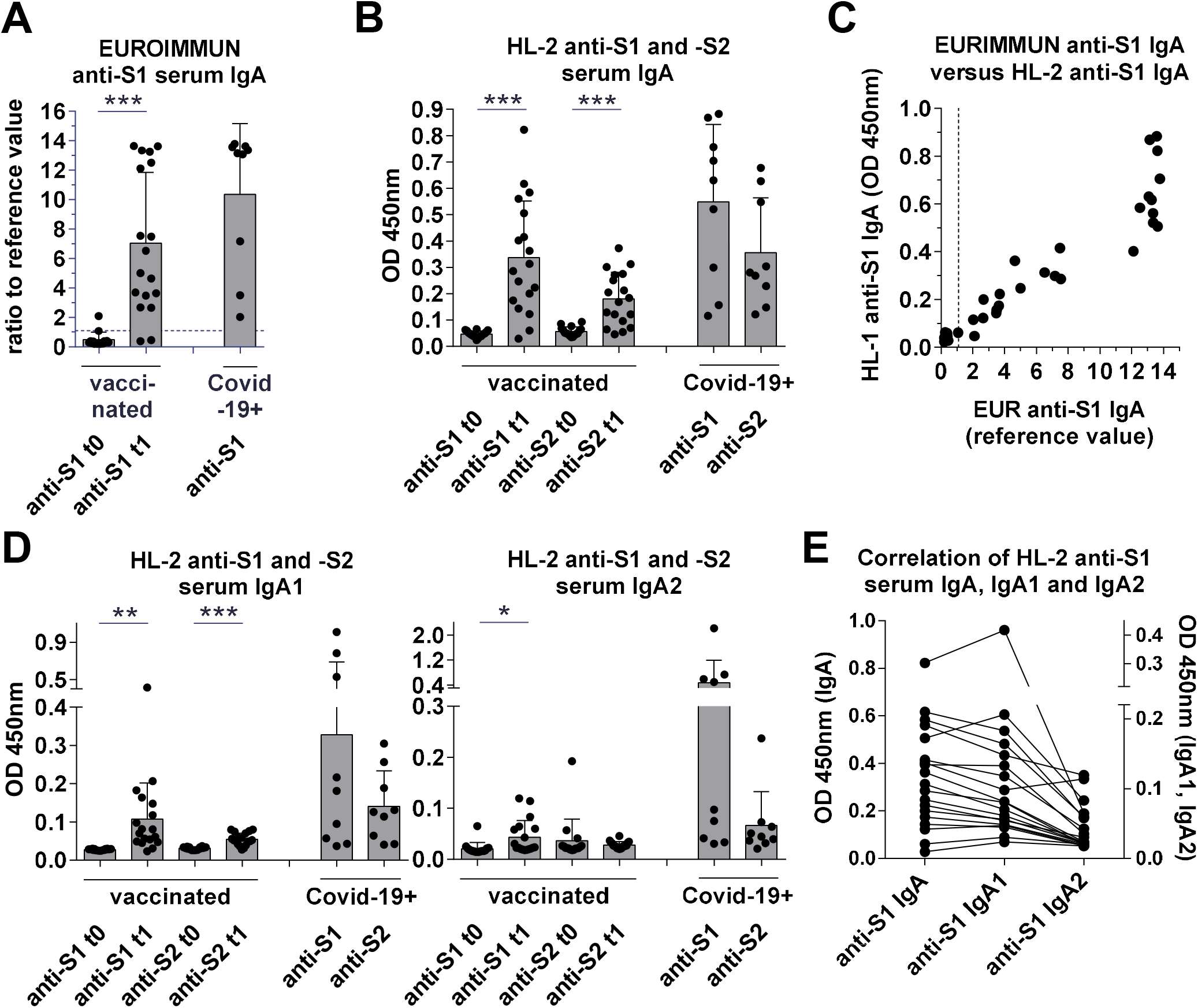
SARS-CoV-2-S1 and -S2 reactive serum IgA, IgA1 and IgA2 Ab ELISA. Sera were collected from vaccinated individuals before and after the first vaccination as well as non-vaccinated individuals with past or current COVID-19 infection and analyzed by anti-SARS-CoV-2-S1 and -S2 IgA and IgA1 and IgA2 ELISA. (**A**) EUROIMMUN anti-SARS-CoV-2-S1 IgA ELISA. (**B**) HL-2 anti-SARS-CoV-2-S1 and -S2 IgA ELISA. (**C**) Correlation between the EUROIMMUN and the HL-2 anti-SARS-CoV-2-S1 IgA ELISA. (**D**) HL-1 anti-SARS-CoV-2-S1 and -S2 IgA1 and IgA2 ELISA. (**E**) Correlation between HL-2 anti-SARS-CoV-2-S1 IgA, IgA1 and IgA2 ELISA results.

To analyze anti-SARS-CoV-2-S1 and -S2 IgA1 and IgA2 Abs, we used well established monoclonal anti-human IgA1 and IgA2 secondary Abs in our HL-2 ELISA system (**Supplementary Fig. 1**). The vaccinated IgA+ individuals developed anti-S1 IgA1 and some individuals, in particular proband no. 10 and no. 15, also IgA2 serum Abs (**Fig. 3D and E and Supplementary Fig. 2**).

### The vaccine induces anti-SARS-CoV-2-S1 IgG Abs in the saliva

Infection of the respiratory tract with SARS-CoV-2 induces anti-SARS-CoV-2 IgA as well as IgG Abs in the saliva (**Isho et al**., **2020**). To investigate the induction of anti-SARS-CoV-2 IgA and IgG Abs in the saliva after vaccination, saliva was collected after the first immunization of 14 vaccinated individuals at the indicated time point as well as from 18 individuals without known COVID-19 history (**Fig. 1 and Supplementary Table 1**) and analyzed by using the HL-2 ELISA protocol. Saliva from COVID-19 infected individuals of the ICU of our hospital were tested positive during establishment of the HL-2 anti-S1 IgA ELISA (data not shown). An enrichment of anti-SARS-CoV-2-S1 IgA Abs could hardly be detected in the saliva of vaccinated as compared to non-vaccinated individuals (**Fig. 4A**). However, vaccinated individuals showed an enrichment of anti-S1 IgG Abs compared to non-vaccinated individuals (**Fig. 4B**). The two lowest anti-S1 IgG values in the vaccinated group belonged again to proband nr. 11 and nr.19. The data show that the success of vaccination can be analyzed by detecting the induction of anti-SARS-CoV-2-S1 IgG Abs in the saliva.

**Fig. 4.**
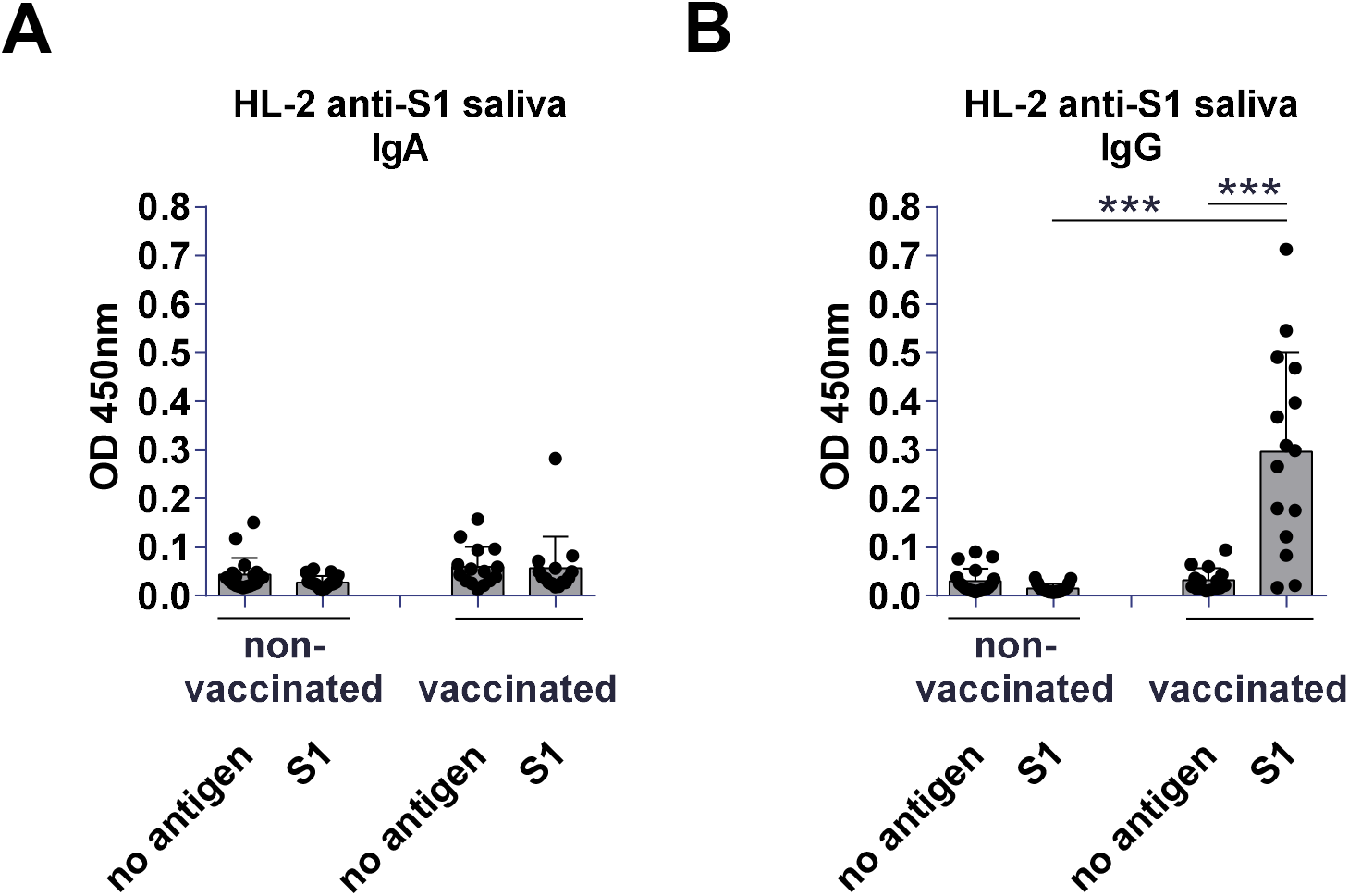
SARS-CoV-2-S1 and -S2 reactive saliva IgG and IgA Ab ELISA. Saliva was collected from vaccinated individuals as well as non-vaccinated individuals without known COVID-19 history and analyzed by HL-2 anti-SARS-CoV-2-S1 (**A**) IgA and (**B**) IgG ELISA.

### The vaccine induces SARS-CoV-2-S1-reactive IgG+ and IgA+ PCs and potential memory B cells

Blood samples of the vaccination study cohort before and after the first immunization and of individuals with past or current COVID-19 infection were collected at the indicated time points and PBMCs were isolated. The PBMCs were stained with fluorochrome-coupled Abs against extracellular surface markers of PCs (that may contain short-lived plasma blasts and long-lived plasma cells) as well as memory B cells (CD19, CD38, IgG, IgA) and intracellular molecules (IgG and IgA) together with biotinylated SARS-CoV-2-S1 antigen and fluorochrome-coupled streptavidin and analyzed by flow cytometry.

Individuals with current COVID-19 infection showed in particular S1-reactive class-switched IgG+ and/or IgA+ PCs (CD19int, CD38hi) and individuals with past COVID-19 infection potential S1-reactive IgG+ and/or IgA+ memory B cells (CD19+, CD38int/(lo)) (**Fig. 5A-H** and data not shown). At time point zero before vaccination, the individuals of the vaccination study cohort showed hardly any S1-reactive B cells (**Fig. 5A and B**). However, after immunization S1-reactive IgG+ PCs and potential memory B cells as well as for several individuals some S1-reactive IgA+ PCs and/or potential memory B cells could be detected (**Fig. 5A-H**).

**Fig. 5.**
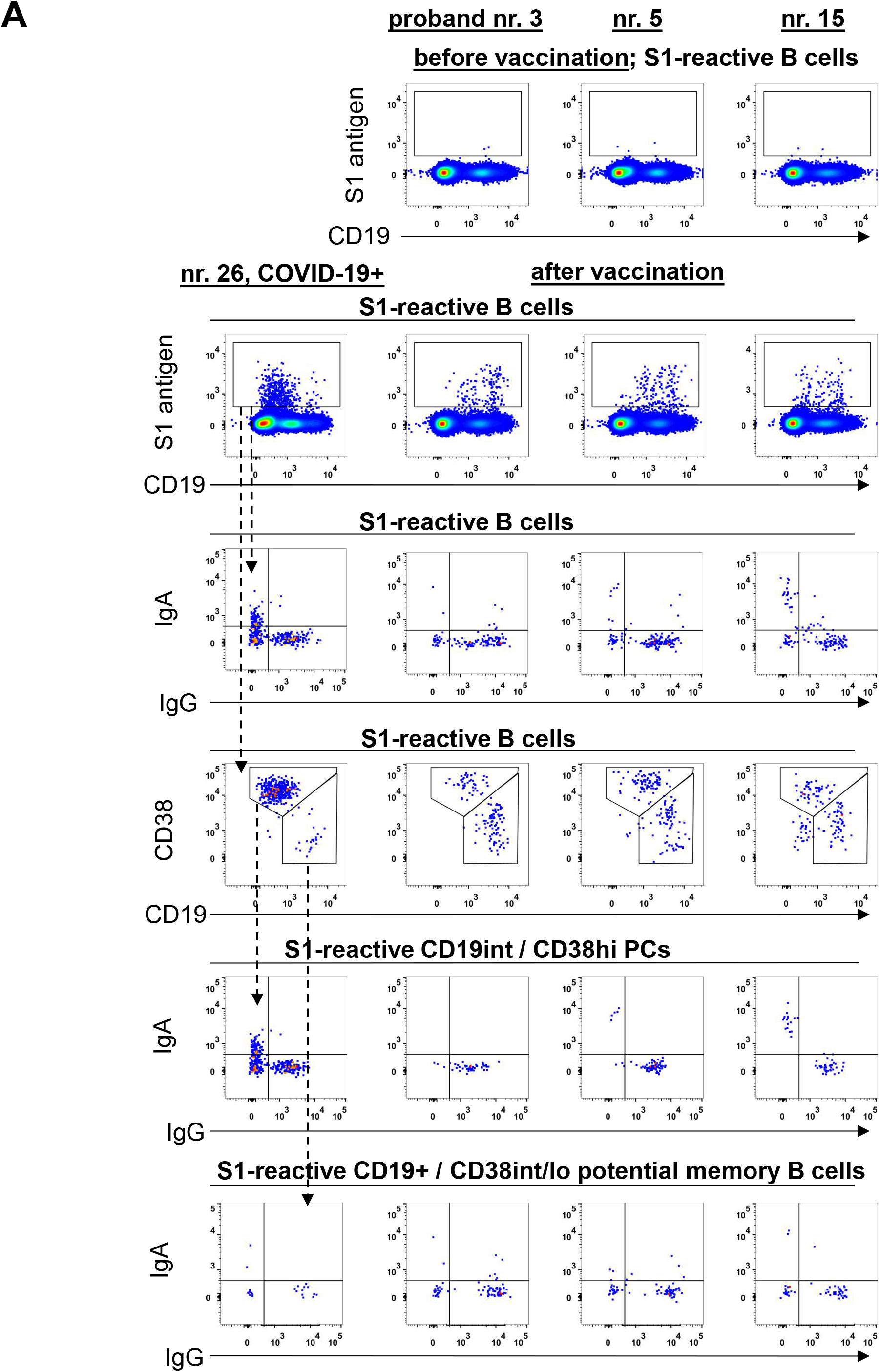
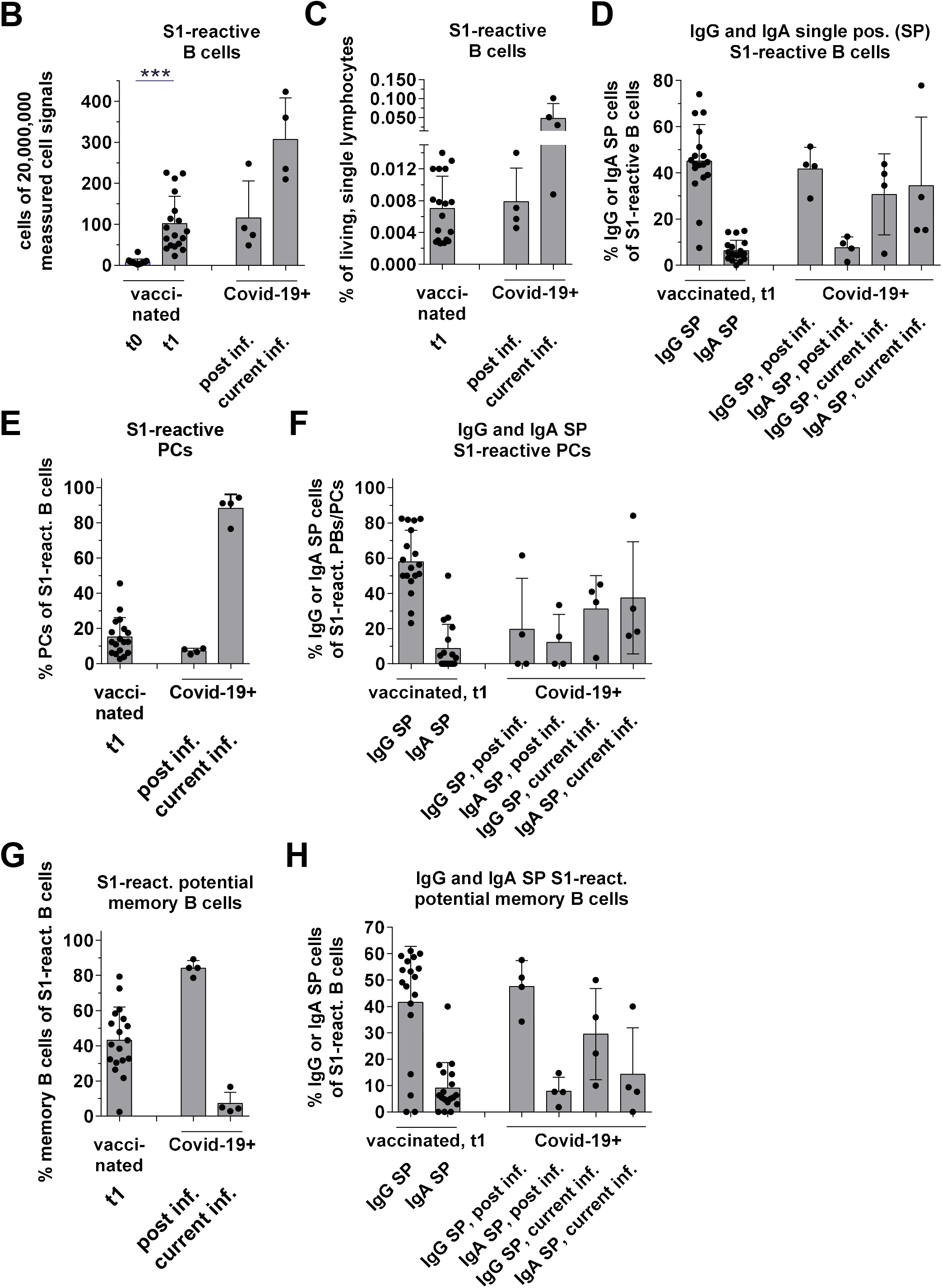
SARS-CoV-2-S1 reactive IgG and IgA blood PCs and potential memory B cells analyzed by flow cytometry. Blood cells from the vaccination study cohort before and after the first immunization as well as from non-vaccinated individuals with past or current COVID-19 infection were analyzed by flow cytometry. Lymphocytes were encircled in the FSC-A/SSC-A analysis and further gated for single (FSC-A/FSC-H) and living cells. These cells were analyzed for SARS-CoV-2-S1-reactive IgG+ and IgA+ (**A-D**) total cells, (**E and F**) PCs (CD19int, CD38hi) and (**G and H**) potential memory B cells (CD19+, CD38int/(lo)). Example analyses of one current COVID-19 ICU patient and three individuals of the study cohort are shown in (A).

## Discussion

Upon SARS-CoV-2 infection, IgM, IgG and IgA Ab responses (Isho et al., 2020) and memory T and B cells have been detected (Chen and John Wherry, 2020; Rodda, 2020). However, studies with other human coronavirus-strains suggest protective immunity to be rather short lasting (Edridge et al., 2020), so the long-term effect and also all different protection mechanisms of the T and B cell and Ab responses against SARS-CoV-2 are still under investigation.

For the BioNTech / Pfizer mRNA vaccine BNT162b2, protection against COVID-19 has also been described for the first weeks after the second vaccination, but long-term effectiveness and protecting mechanisms are unclear as well. In both cases, defense mechanisms of neutralizing Abs have been described. However, the protective immune response upon vaccination against SARS-CoV-2 might differ from the pathogen-induced immune response. Vaccine-induced immune responses are in particular dependent on T cell help, the type of adjuvant effect and the application route.

After the single vaccination with BNT162b2, mainly anti-SARS-CoV-2-S1- and -S2 serum IgG1 and IgG3 Abs could be detected. Furthermore, IgG+ PCs and potential memory B cells were observed as described before (Wang et al., 2021). Both observations suggest that the vaccine induces a proper CD4+ T cell-dependent immune response with memory B cell formation. The adjuvant effect of the lipid nano-particles containing the mRNAs is, however, unclear. Different adjuvant effects might modulate the induction of distinct IgG as well as IgA subclasses in the serum. Besides IgG1 and IgG3 as well as IgG2 in some subjects (nr. 3 and nr. 6), IgG4 was not detected. In addition, the vaccine induced anti-SARS-CoV-1-S1 serum IgA1 and in some individuals, in particular in proband nr. 10 and nr. 15, also IgA2 serum Abs.

Besides Fab-part-mediated neutralizing mechanisms of Abs, Fc-part mediated effector mechanisms can include the recruitment of immune cells through binding to Fcgamma receptors as well as the activation of the complement system for fighting against the virus. Both, IgG1 and IgG3 show the highest affinities for activating Fcgamma receptors (Lux and Nimmerjahn, 2013), whereas IgG4, that cannot activate complement, is often linked to less powerful and also anti-inflammatory effector functions by for example inhibiting IgG1 and IgG3 effector mechanisms or preferentially interacting with the inhibitory Fcgamma receptor IIB (Lilienthal et al., 2018).

Mucosal barriers represent the site of entry and at the same time the first obstacle for the virus. It is conceivable that the mucosa is the earliest possible place to stop the SARS-CoV-2 infection and mucosal Abs might play a crucial role. SARS-CoV-2 infection induces a broad IgM, IgA and IgG response in the serum as well as in the saliva (Isho et al., 2020).

For vaccination against mucosal pathogens different approaches are followed to induce mucosal first line of defense immune responses. Intranasal vaccination, boost injections into the mucosa, specific adjuvants and the induction of tissue resident CD4+ memory T cells for fast mucosal immune responses after pathogen contact are considered (Bartsch et al., 2020; Krammer, 2020).

Although anti-S1 IgA could hardly be detected in the saliva, the existence of S1-reactive class switched IgA+ PCs and anti-S1 serum IgA1 and in some individuals also IgA2 might be precursors for rapid secretory IgA1 and IgA2 responses in the respiratory tract after pathogen contact. Individuals with anti-S1 serum IgA1 and/or IgA2 might produce secretory IgA1 and IgA2 faster than individuals without anti-S1 serum IgA1 and IgA2.

However, while ant-S1 IgA could hardly be detected in the saliva, anti-S1 IgG was observed after vaccination in the saliva and might already present certain protection against SARS-CoV-2 in particular in the lower part of the respiratory tract (Krammer, 2020).

Thus, these general and also individually specific immune responses might contribute to first Ab defense lines against SARS-CoV-2 and also its mutations (Arif, 2021).

## Data Availability

Our ethics does not allow to share the data without signing an agreement. Parties interested can contact the corresponding author to obtain access to this data.

## Acknowledgments

We thank Andreas Recke and Ralf Ludwig for the monoclonal anti-collagen type VII IgG1-4, IgA1 and IgA2 Abs. M.E. was funded by the Volkswagen Foundation (97301), the Deutsche Forschungsgemeinschaft (DFG, German Research Foundation) – 398859914 (EH 221/10-1); 400912066 (EH 221/11-1); and 390884018 (Germany’s Excellence Strategies - EXC 2167, Precision Medicine in Chronic Inflammation [PMI]) and the Federal State Schleswig-Holstein, Germany („COVID-19 Research Initiative Schleswig-Holstein”; DOI4-Nr. 3).

## Author contributions

A.S.L. and J.R. established and conducted flow cytometry analyses. I.K., E.L.M., A.S.L., S.L. and J.R. established and performed serum and saliva analyzes. V.v.K., H.B.L., J.S.B., J.L.Q. and M.S. performed further blood and serum characterizations. T.G. organized the blood sampling. J.R. and M.E. performed statistical analysis. J.R. and M.E. had full access to all of the data in the study and take responsibility for the integrity of the data and the accuracy of the data analysis. J.R. and M.E. coordinated and supervised the experiments and wrote the manuscript. All authors verified and improved the manuscript.

## Supplementary Data

**Supplementary Table 1. Subjects and time points of sample collection**

Individuals of the vaccination study cohort as well as negative and positive control individuals and the time points of sampling.

**Supplementary Fig. 1. Specificity control of anti-IgG, IgG1-4, IgA, IgA1 and IgA2 secondary Abs**.

ELISA plate wells were coated with monoclonal human IgG1-4, IgA1 and IgA2 Abs and detected with the indicated secondary Abs.

**Supplementary Fig. 2. Correlation between anti-SARS-CoV-2-S1 IgG, IgG subclass, IgA and IgA subclass Ab ELISA results shown in Figs. 2 and 3**.

