## Supplementary figures and images for "The BioNTech / Pfizer vaccine BNT162b2 induces class-switched SARS-CoV-2-specific plasma cells and potential memory B cells as well as IgG and IgA serum and IgG saliva antibodies upon the first immunization"

### Suppl. Figures 1+2

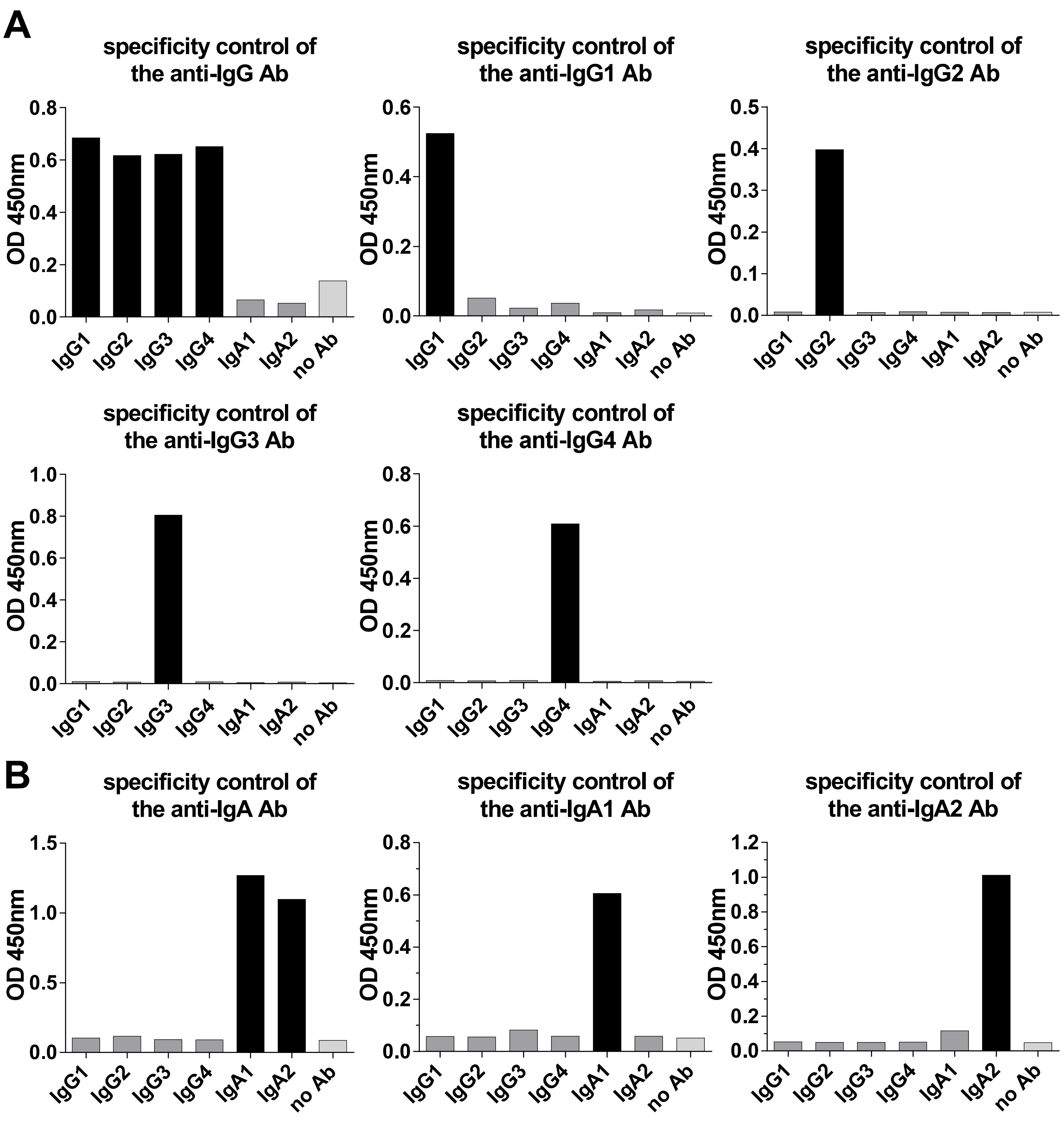

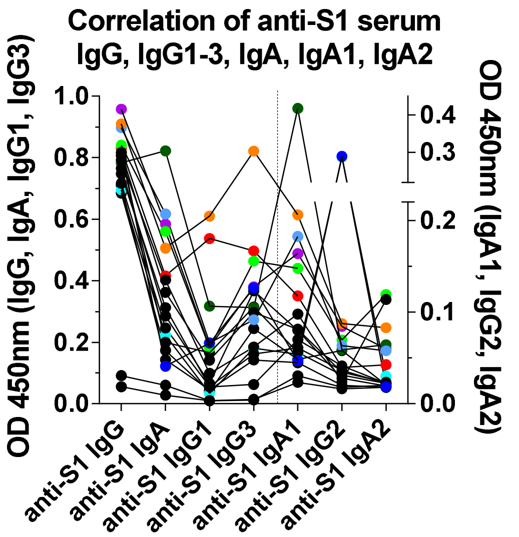
